# Distinct temporal dynamics of motor and neuropsychiatric responses to levodopa in Parkinson’s disease

**DOI:** 10.64898/2026.05.22.26353856

**Authors:** Damien Benis, Sabina Catalano Chiuvé, Charline Rime, Christo Bratanov, Julien F Bally, Vanessa Fleury

## Abstract

**Background:** Neuropsychiatric fluctuations in Parkinson’s disease (PD) often accompany motor fluctuations, but their temporal relationship during the acute levodopa response remains unclear.

**Objectives:** To determine whether motor and neuropsychiatric responses occur synchronously during the OFF-to-ON transition.

**Methods:** Nineteen fluctuating PD patients underwent a high-resolution levodopa challenge with repeated assessments every 10 minutes for 70 minutes after levodopa administration. Motor symptoms (akinesia, rigidity) and neuropsychiatric fluctuations were quantified. Transition times (t25%-t50%-t75%-t100%) and response profiles were analyzed using correlation and clustering approaches.

**Results:** Motor and neuropsychiatric transition times were not correlated at any threshold (all FDR-corrected p>0.05; Bayes factors <1), supporting temporal dissociation. Among 18 patients with complete data, clustering revealed synchronous (6/18), neuropsychiatric-preceding (7/18), and motor-preceding (3/18) profiles.

**Conclusion:** Motor and neuropsychiatric responses to levodopa during PD fluctuations are partly independent and follow heterogeneous, patient-specific temporal profiles, supporting the search for distinct biomarkers and future individualized adaptative therapies.

## Introduction

Neuropsychiatric fluctuations (NPF) in Parkinson’s disease (PD) are transient changes in mood, inner tension, motivation, sense of well-being, or self-confidence, occurring in relation to dopaminergic treatment cycles (“ON-OFF” states).^1^ They constitute a core component of nonmotor fluctuations (NMF), and are often highly disabling.^2–4^

Longitudinal data indicate that NMFs typically emerge concomitantly with motor fluctuations (MFs) or within months after their onset, and only rarely precede them.^5^ This supports a close but nonparallel temporal relationship at the disease level. At a shorter timescale (days), studies specifically examining NPF have shown that their temporal relationship with MF is only partial, suggesting partly distinct underlying mechanisms.^6–8^ However, the precise time course of MF and NPF symptoms in the acute 1-hour period of the levodopa challenge has not yet been investigated.

The levodopa challenge provides a standardized model to study short-term dopaminergic effects across the transition from clinical OFF to ON states.^9^ The recent multicenter validation of the Neuropsychiatric Fluctuation Scale provides a robust and disease-specific tool to assess OFF- and ON-related neuropsychiatric symptoms in real time, with good psychometric properties.^10^

In the present study, we investigated the temporal evolution of neuropsychiatric and motor symptoms during the first 70 minutes of a levodopa challenge in PD patients with fluctuations. We specifically tested whether neuropsychiatric responses emerge synchronously with motor improvement or follow a distinct temporal profile, which would support a partial dissociation between motor and non-motor dopaminergic responses.

## Methods

This study was approved by the Geneva Ethics Committee (EC-2021-02341). All participants provided written informed consent.

### Participants

Patients were recruited from the Movement Disorders Unit of the Geneva University Hospital and the Lausanne University Hospital. Inclusion criteria were PD^11^ treated with dopaminergic replacement therapy (DRT) including levodopa, and the presence of MF and NPF defined respectively by a score ≥1 on item 4.1 and/or on item 4.3 of the Movement Disorder Society-Unified PD Rating Scale (MDS-UPDRS) IV^12^, and a score ≥1/4 on item III of the Ardouin Scale of Behavior in Parkinson’s Disease (ASBPD^13^). Exclusion criteria included >80 years and cognitive impairment (Montreal Cognitive Assessment MOCA^14^ <25/30).

### Procedure

Patients completed a baseline visit under regular DRT during which demographic and clinical characteristics were recorded: disease duration, Hoehn and Yahr (H&Y) stage^15^, levodopa-equivalent daily dose (LEDD)^16^, motor severity (MDS-UPDRS-III)^12^, non-motor symptoms (PD-NMS)^17^, behavioral assessment (ASBPD)^13^, depression (BDI)^18^, and quality of life (PDQ-8)^19^.

During a second visit, a high-resolution levodopa challenge was performed. Patients were assessed after overnight withdrawal from dopaminergic medication (OFF), in a fasting state. They received 150% of their usual morning levodopa dose (levodopa-benserazide dispersible formulation) to ensure rapid and standardized dopaminergic stimulation. Motor function (MDS-UPDRS III^12^ and Marconi dyskinesia scale^20^) and neuropsychiatric symptoms (Neuropsychiatric Fluctuation Scale (NFS)^10^, Supplementary Information (SI)-1; State-Trait Anxiety Inventory for Adults^21^ (STAI) and the Hospital Anxiety and Depression Scale (HAD)^22^) were evaluated in the OFF state and at peak levodopa dose.

To capture the transition from OFF to ON states, abbreviated 32-point version of the MDS-UPDRS III (akinesia measurement with items 3.4 and 3.7, rigidity with items 3.3 upper and lower limbs) and the full NFS (including ON- and OFF-related sub-scores rated simultaneously, independently of the patient’s current dopaminergic state) administered on a tablet, were repeated every 10±5 minutes during the 70 minutes after levodopa intake. This high-temporal-resolution design allowed characterization of the dynamic evolution of motor and neuropsychiatric responses to levodopa, rather than their comparison at static OFF and peak ON states (SI-2).

### Data analysis

Analyses were performed using Python and JASP. *P*-values were corrected for multiple comparisons using the false discovery rate (FDR).^23^ Clinically meaningful fluctuations were defined as ≥4 points on the NFS-ON and/or NFS-OFF subscores for NPF and a change of ≥1 point on the abbreviated akinesia score (range 0-4) for MF. Patients below these thresholds were excluded from transition analyses (SI-3).

The “transition phase” was defined as the 70 minutes following levodopa intake. Transition times (t25%, t50%, t75%, t100%) for motor and non-motor responses were defined as the time to reach 25%, 50%, 75%, and 100% of the maximal improvement following levodopa intake (SI-4).

Associations between motor and neuropsychiatric transition times were assessed using both frequentist and Bayesian correlation analyses (with NFS-ON and NFS-OFF sub-scores analyzed separately). Bayes factors (BF) above 1 supports the association between the scores, while a BF below 1 favors the absence of association between the scores (SI-3).^24^

Hierarchical DBCAN clustering based on t50% and t100% identified distinct temporal response profiles (SI-4, SI-5).^25^

## Results

Nineteen patients were included. Rigidity scores were available for 15 patients. Overall, patients had moderate disease severity, preserved cognition, and minimal depressive symptoms (SI-6) as well as motor and neuropsychiatric fluctuations with a significant peak ON-state improvement in motor severity and neuropsychiatric symptoms (SI-7).

### Temporal dissociation between motor and neuropsychiatric responses during the OFF-to-ON transition

We first investigated the temporal relationship between motor (akinesia or rigidity) and neuropsychiatric responses (NFS-ON and NFS-OFF subscores) during the OFF-to-ON transition using multiple correlation analysis on timesteps with partial sample sizes due to missing parameter estimates (one patient exhibited significant NFS-ON neuropsychiatric fluctuations, without significant NFS-OFF neuropsychiatric fluctuations, and one patient had a fluctuation pattern precluding akinesia t25% estimation).

No significant temporal correlations were observed between motor (akinesia or rigidity) and neuropsychiatric transition times at either threshold (t25%, t50%, t75% and t100%) (all FDR-corrected, p>0.05, Fig.1, SI-8). Bayesian analyses consistently supported the absence of association, with BF predominantly being ∼0.50 across timepoints (<0.33 for NFS-OFF vs Akinesia from t25% to t75%, NFS-OFF vs Rigidity at t25%, and for NFS-ON vs akinesia at t100%), in favor of independence between motor and neuropsychiatric dynamics. Detailed transition curves and derived timepoints are provided in the SI-9-10.

**Figure 1.**
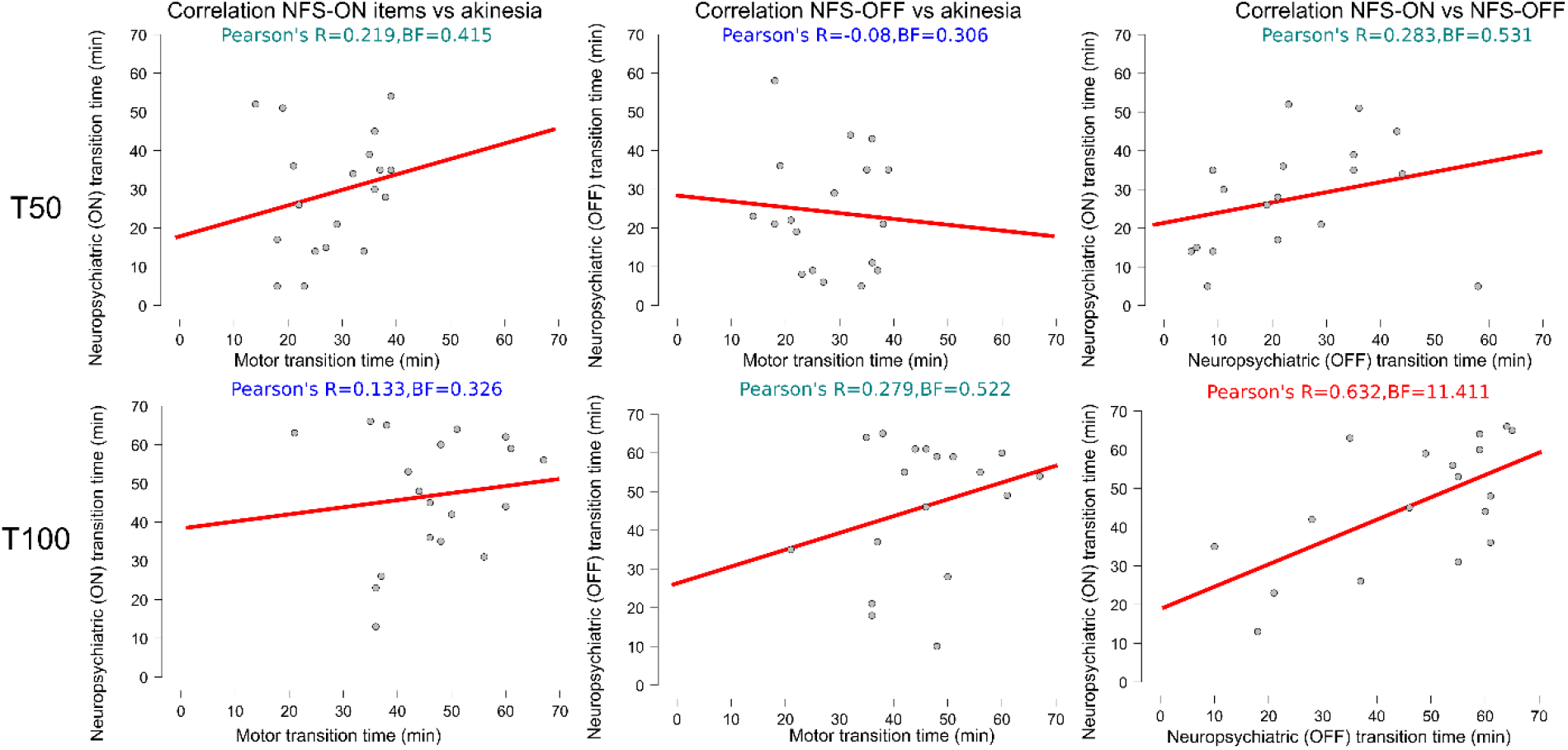
Lack of temporal coupling between motor and neuropsychiatric responses to levodopa. Each point represents one patient. Scatterplots illustrate the relationship between motor (akinesia) and neuropsychiatric (NFS-OFF items, NFS-ON items) transition times at two response thresholds: t50% (upper panels) and t100% (lower panels). No significant association was observed between akinesia and either NFS-ON or NFS-OFF transition times at any threshold after FDR correction, supporting a temporal dissociation between motor and neuropsychiatric responses. In contrast, NFS-ON and NFS-OFF transition times were positively correlated at t100%, suggesting convergence of neuropsychiatric ON and OFF dynamics at later stages of the levodopa response. Pearson correlation coefficients (R) and Bayes factors (BF) are indicated in each panel.

In contrast, OFF and ON neuropsychiatric transition times presented evidence of correlation (BF=11.41) at the t100% threshold (Fig.1), but not at early timepoints (t25%, t50%), suggesting a late-stage partial convergence of ON and OFF neuropsychiatric states, although *p*-value did not survive FDR correction (p=0.14).

Overall, these findings indicate a temporal dissociation between motor and neuropsychiatric responses to levodopa.

### Heterogeneity of motor and neuropsychiatric temporal response patterns

To further explore interindividual variability, we performed an unsupervised clustering analysis based on transition times in the 18 patients with complete datasets (excluding the patient with no NFS-OFF fluctuations). Three main patterns of motor-neuropsychiatric dynamics emerged. A cluster example (subgroup) for each pattern is presented in Figure 2. Mean transition times, individual patterns and average patterns for all clusters are presented in SI-9-10-11-12.

**Figure 2.**
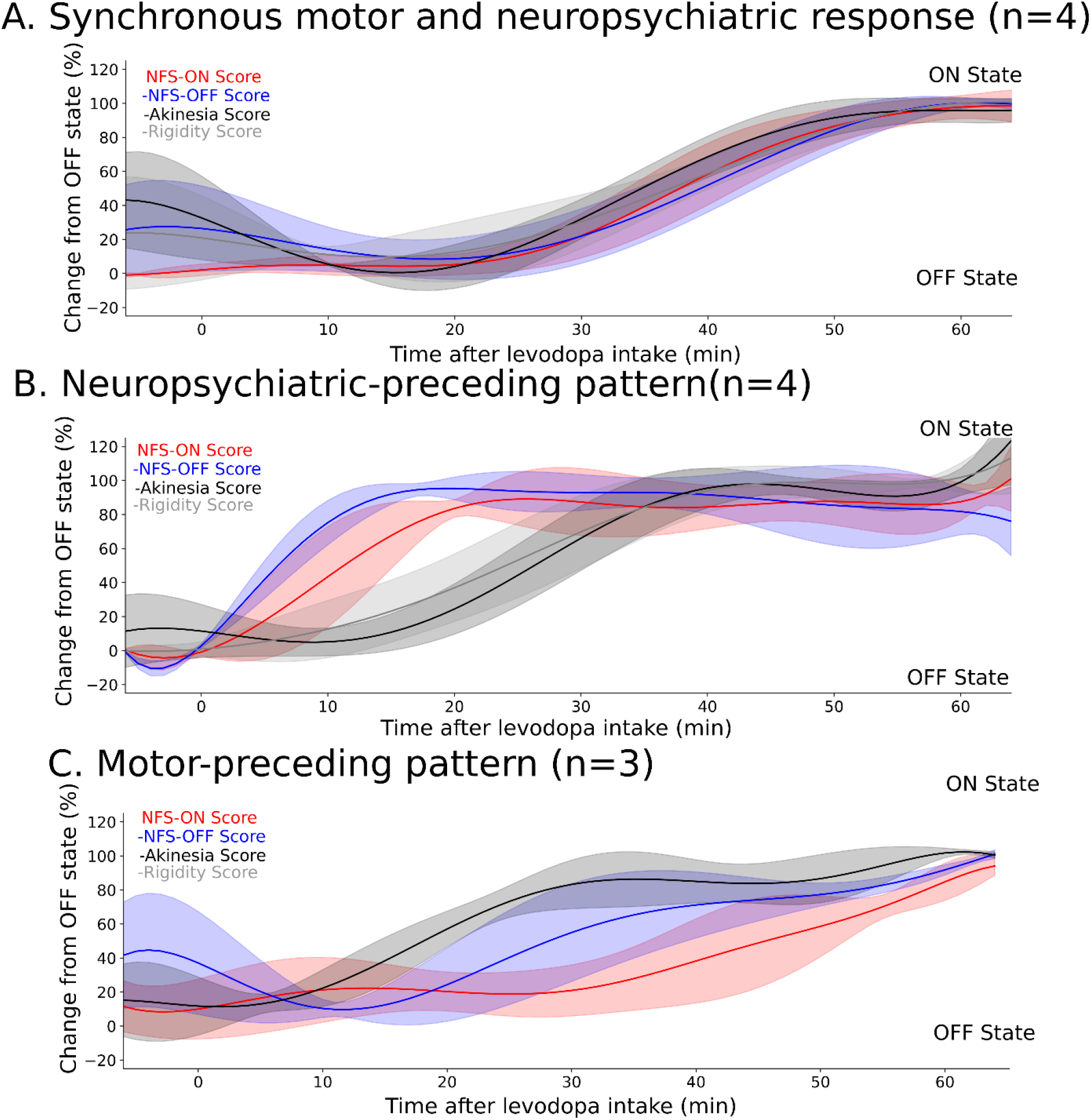
Three temporal patterns of motor and neuropsychiatric responses during the OFF-to-ON transition. Mean normalized trajectories of neuropsychiatric and motor scores during the 60 minutes following levodopa intake, grouped according to unsupervised clustering of transition times. Three main patterns were identified: synchronous transitions, earlier neuropsychiatric changes, and earlier motor responses. The most representative cluster for this pattern has been selected for the figure. Curves represent mean values, with shaded areas indicating standard deviations. OFF-related neuropsychiatric scores are inverted for visualization.

First, a synchronous pattern was observed in 6 patients, in whom motor and neuropsychiatric responses evolved in parallel. This included an early-synchronous subgroup (n=4, Fig 2-A, t50% around 24-26 min) and a delayed-synchronous subgroup (t50% around 37 min) (n=2, SI-9-10-11-12).

Second, a neuropsychiatric-preceding pattern was identified in 7 patients, characterized by earlier changes in neuropsychiatric symptoms compared to motor improvement. In one subgroup (n=4), both ON- and OFF-related neuropsychiatric symptoms improved early (t50% around 11-16 min) (Fig 2-B, SI-9-10-11-12), while motor improvement occurred later (t50% around 28 min). In another subgroup (n=3), OFF-related neuropsychiatric symptoms improved early (t50% around 14 min), whereas motor and ON-related neuropsychiatric symptoms followed later (t50% ≈ 31– 37 min) (SI-9-10-11-12).

Third, a motor-preceding pattern was observed in 3 patients, in whom motor responses occurred earlier (t50% around 18 min) than neuropsychiatric changes (t50% around 27-46 min) (Fig 2-C, SI-9-10-11-12).

Overall, these findings reveal marked interindividual heterogeneity, with distinct temporal profiles ranging from synchronous to dissociated motor and neuropsychiatric responses. Additional exploratory associations between transition delays and clinical profiles are reported in SI-9 and SI-11.

## Discussion

Using repeated assessments every 10 minutes during the first 70 minutes after levodopa intake, we demonstrated a temporal dissociation between motor and neuropsychiatric effects of levodopa in fluctuating PD patients, together with marked interindividual variability, supporting patient-specific temporal profiles of dopaminergic effect.

The temporal dissociation between motor and neuropsychiatric improvement following levodopa administration suggests that the mechanisms underlying motor and neuropsychiatric fluctuations may be partly independent. This is consistent with previous studies showing that NMF are not systematically aligned with MF.^7,26,27^ This interpretation is further supported by longitudinal studies reporting a lack of correlation between motor status and mood variations^28^, as well as distinct temporal profiles, with non-motor OFF periods often lasting longer than motor OFF states.^6^ Most of these studies relied on broad non-motor symptom scales aggregating heterogeneous domains and potentially masking domain-specific dynamics. While a previous ecological study reported a correlation between bradykinesia and OFF-related NPF over several days^7^, our findings indicate that this relationship breaks down at finer temporal resolution. The lack of correlation during the acute OFF-to-ON transition suggests that motor and neuropsychiatric responses may appear coupled at the clinical-state level yet dissociate during rapid dopaminergic dynamics.

Our results reveal distinct temporal dynamics between the resolution of OFF-related neuropsychiatric symptoms and the emergence of ON-related neuropsychiatric features. Although both converged toward maximal ON-state expression at t100%, these processes were not strictly synchronous. While previous studies reported an inverse correlation between ON and OFF-NFS subscores^7,27^, our findings indicate early dissociations between the decline of OFF neuropsychiatric symptoms and the rise of ON features, again suggesting partially distinct neural mechanisms.

Finally, our protocol highlighted marked interindividual variability in temporal response profiles. Three main patterns emerged: motor-preceding, neuropsychiatric-preceding and synchronous transitions. While variability in the timing and expression of NMF has previously been reported^6,26,27^, prior studies were not designed to capture the fine temporal dynamics of acute levodopa responses. To our knowledge, our study is the first to address such variability during the acute OFF-to-ON transition. These distinct profiles may reflect interindividual differences in the degree and topography of dopaminergic denervation across motor and non-motor circuits, including nigrostriatal and mesocorticolimbic pathways, leading to differential temporal responses to levodopa.

The main limitations of this study are the small sample size, the demanding protocol and the subjective nature of the self-reported NFS.

Altogether, this study suggests that motor and neuropsychiatric responses to levodopa are partially independent and follow patient-specific transition profiles. This dissociation provides a rationale for identifying distinct biomarkers of motor and neuropsychiatric states using high-resolution electrophysiological or imaging approaches. Identifying such biomarkers could ultimately enable individualized circuit-based therapeutic strategies, including adaptive closed-loop deep brain stimulation targeting not only motor symptoms, but also neuropsychiatric fluctuations through refined targeting of motor and non-motor dopaminergic networks.

## Data Availability

All data produced in the present study are available upon reasonable request to the authors

## Abbreviations

ASBPD: Ardouin Scale of Behavior in Parkinson’s Disease
BDI: Beck Depression Inventory
BF: Bayes Factor
DRT: dopamine replacement therapy
FDR: false discovery rate
HAD: Hospital Anxiety and Depression scale
H&Y: Hoehn and Yahr
LEDD: levodopa equivalent daily dose
MDS-UPDRS: Movement Disorder Society-Unified Parkinson’s Disease Rating Scale
MF: motor fluctuations
MOCA: Montreal cognitive assessment
NFS: Neuropsychiatric fluctuations scale
NMF: non-motor fluctuation
NMS: non-motor symptoms
NPF: Neuropsychiatric fluctuations
PD: Parkinson’s disease
PDQ-8: Parkinson’s disease Questionnaire-8 items
SI: Supplementary Information
STAI: State-Trait Anxiety Inventory for Adult.

## Acknowledgments

We cordially thank all participants for having taken part in this study. We thank the residents Natalia Drobinska, Damien Schneider and Parkinson’s nurses Emilie Tomkova, Nathalie Hauri and Joris Callot for their valuable assistance in conducting the levodopa challenge assessments in patients. We thank Giannina Iannotti for her assistance with study organization, including facilitating coordination with the center where the experiments were conducted. We thank the Swiss Foundation for Innovation and Training in Surgery for granting access to their facilities.

## Authors’ Roles

1. Research project: A. Conception, B. Organization, C. Execution;
2. Statistical Analysis: A. Design, B. Execution, C. Review and Critique;
3. Manuscript Preparation: A. Writing of the first draft, B. Review and Critique;

DB: 1A, 1C, 2A, 2B, 3A

SCC: 1A, 1B, 1C, 2C, 3B CB: 3B

CR: 1A, 1C, 3B

JFB : 3B

VF: 1A, 1B, 2C, 3B

## Disclosures

### Financial Disclosures for the previous 12 months

Dr.med. Vanessa Fleury reports having received travel grants from Insightec, Bial, and Merz to attend scientific meetings. These supports are unrelated to the present study. Dr.med Julien F Bally reports having received travel grants from Merz, Abbvie, Medtronic, Spirig, Bial, Zambon to attend scientific meetings and payment for lectures or advisory boards.

The other authors declare that there are no additional disclosures to report.

