## Supplementary Information (SI) for "Distinct temporal dynamics of motor and neuropsychiatric responses to levodopa in Parkinson’s disease"

**SUPPLEMENTARY DATA**

### 1. Description of the Neuropsychiatric fluctuation scale (NFS)

The NFS is a 20-item self-report questionnaire specifically developed to assesses neuropsychiatric fluctuations in Parkinson’s disease patients. It provides separate ON- and OFF-related sub-scores, each comprising 10 items. ON-NFS items capture symptoms typically associated with dopaminergic stimulation (e.g., well-being, increased confidence, increased energy), whereas OFF items assess symptoms related to dopaminergic withdrawal (e.g., depression, anxiety, apathy, fatigue, unease, slowness of thought).

Both ON and OFF sub-scores (NFS-ON items and NFS-OFF items) are rated simultaneously, independently of the patient’s current dopaminergic state, allowing a comprehensive assessment of fluctuation patterns. During the OFF phase of a neuropsychiatric fluctuation, OFF-item scores are typically elevated whereas ON-item scores remain low. Conversely, during the ON phase of a neuropsychiatric fluctuation, ON-item scores become elevated while OFF-item scores decrease.

Each item is scored on a 4-point Likert scale (0–3), yielding a maximum score of 30 for each subscale. The NFS has demonstrated good psychometric properties, including reliability and construct validity, in both its initial development and subsequent multicenter validation studies (1,2).

### 2. ****Study**** ****design and temporal assessment of motor and neuropsychiatric responses during a levodopa challenge: Supplementary Figure 1****

Patients were assessed in the OFF state, then every 10 minutes over 60 minutes following levodopa intake to capture the clinical transition to ON state. Motor and neuropsychiatric symptoms were recorded at each timepoint, allowing comparison of their temporal dynamics.

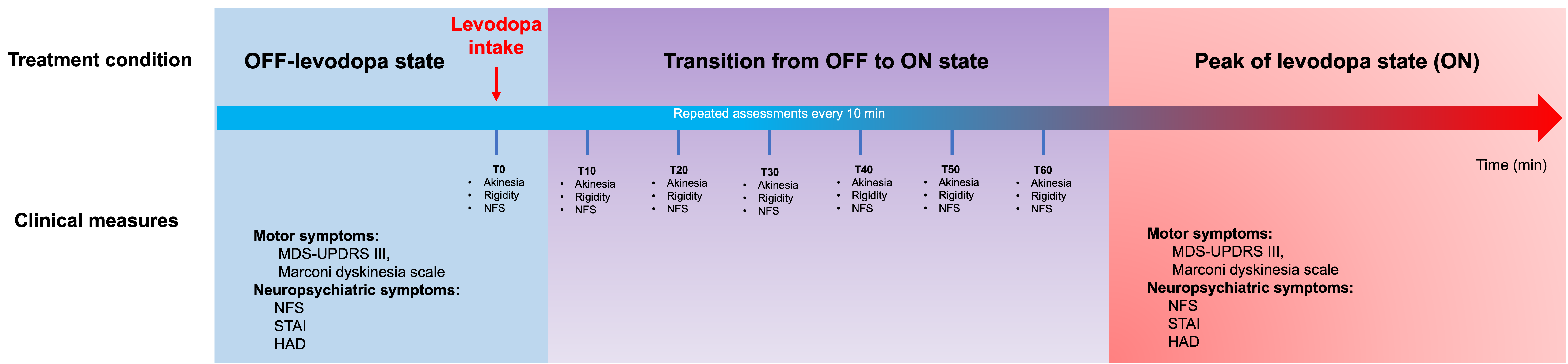

### 3. Definition of transition analysis and preprocessing

*Selection of meaningful fluctuations*

To exclude spurious variations and retain clinically relevant changes, transition analyses were restricted to patients exhibiting meaningful motor and neuropsychiatric fluctuations. Meaningful fluctuations were defined as a ≥4-point change in NFS ON or OFF sub-scores (range 0-30), and a ≥1-point change in akinesia or rigidity scores (range 0-4), between the maximal OFF-state and maximal ON-state values observed during the session. These thresholds were chosen to reflect clinically meaningful changes relative to scale ranges and based on typical ON–OFF variation reported in previous studies (1).

*Data normalization and interpolation*

The “transition phase” was defined as the 60 minutes following levodopa intake.

For each patient, motor (akinesia, rigidity) and neuropsychiatric (NFS-ON and NFS-OFF) scores were expressed as relative changes from the maximal OFF-state value:

Δscore(t) = score(t) – score_OFF max_

This normalization allowed comparison of transition dynamics across patients.

To obtain continuous transition curves, time series were interpolated using B-spline functions (scipy.interpolate), with values estimated at 1-minute intervals from levodopa intake to 65 minutes (5-knot spline).

##### ***Definition of transition timepoints***

##### Four transition timepoints were computed for each score: **t25**, **t50**, **t75**, and **t100**, defined as the earliest time at which the observed or interpolated signal reached 25%, 50%, 75%, and 100% of the maximal response recorded during the session.

##### ***Statistical analysis of transition dynamics***

##### Transition timepoints were used to assess relationships between motor and neuropsychiatric dynamics using both frequentist and Bayesian correlation analyses (JASP software). Bayes factors (BF) were used to quantify evidence for or against an association. BF > 3: moderate evidence for association. BF < 1/3: moderate evidence for absence of association. This approach allowed formal testing of both the presence and absence of relationships between motor and non-motor transition dynamics (3).

### 4. Clustering pipeline validation

To further characterize the transition profiles observed in our patient’s pool, t50 and t100 timepoints for akinesia and non-motor NFS scores were entered as column (patient identity as line) into a Hierarchical DBCAN clustering algorithm with a minimal cluster size of 2. This method was chosen for its unsupervised cluster number selection, outlier handling, and robustness with high dimensional data (McInnes et al., 2017).

Clustering analysis was limited to t50 and t100 to limit the dimensionality of the clustering space while preserving a meaningful characterization of the temporal dynamics. This selection was guided by two considerations. First, t25 was excluded due to a non-estimable value in one patient, which would have introduced missing data into the analysis. Second, t75 was discarded on the grounds of substantial redundancy with t50, providing no additional discriminative information.

Clustering robustness was assessed using multiple distance metrics (Euclidean, Manhattan, Bray–Curtis). Euclidean and Manhattan distances yielded identical clustering solutions, whereas Bray-Curtis distance resulted in a lower clustering quality (Calinski-Harabasz index decrease of 6.43) and was therefore not retained. Cluster assignment reliability was high, with a mean membership probability >0.80. Fourteen out of 18 patients had a probability >0.80, and 16 out of 18 above 0.60.

### 5. Unclassified patients

Two patients were not assigned to any cluster and were considered outliers. These patients exhibited atypical transition patterns. One patient showed early motor and NFS-ON transitions (t50 around 5 and 15 min), but a markedly delayed NFS-OFF transition (t50 around 58 min). The other showed an early and synchronous transition across all measures (t50 ≈ 17–21 min).

### 6. Demographic and clinical characteristics: Supplementary Table 1

Demographic and clinical characteristics recorded under regular DRT: disease duration, Hoehn and Yahr (H&Y) stage, levodopa-equivalent daily dose (LEDD), motor severity (MDS-UPDRS-III), non-motor symptoms (PD-NMS), behavioral assessment (ASBPD), depression (BDI), and quality of life (PDQ-8)

##### Values are reported as mean ± standard deviation (SD).

| **Variables** | **Value (Mean ± SD or n)** |
| --- | --- |
| Number (Male / Female) | 8 / 11 |
| Age (years) | 65.5 ± 8.2 |
| Disease duration (years) | 10.56 ± 3.79 |
| Most affected side (Left/Right) | 14 / 5 |
| Hoehn and Yahr stage | 2.4 ± 0.4 |
| LEDD (mg/day) | 1071 ± 534 |
| PD-NMS (/30) | 9.8 ± 5.1 |
| BDI (/63) | 9.3 ± 5.5 |
| MoCA (/30) | 27.9 ± 1.4 |
| PDQ-8 (/32) | 9.0 ± 3.9 |

### 7. Changes in motor and neuropsychiatric scores between OFF and peak of dose ON levodopa states during the levodopa challenge: Supplementary Table 2

Values are expressed as mean ± standard deviation (SD). Comparisons between OFF and ON states were performed using Wilcoxon signed-rank tests.

Bonferroni correction was applied for multiple comparisons (significance threshold p < 0.008).

*** indicates statistically significant differences after correction; ns = not significant.

Positive z-values indicate a decrease from OFF to ON, whereas negative z-values indicate an increase.

| **Variable** | **OFF-state**  **(mean ± SD)** | **ON-state**  **(mean ±SD)** | **z** | **p** | **Bonferroni significance (p<0.008)** |
| --- | --- | --- | --- | --- | --- |
| NFS OFF items (/30) | 13.68 ± 7.77 | 3.89 ± 3.64 | 3.621 | 0.001 | *** |
| NFS ON items (/30) | 8.68 ± 5.55 | 19.63 ±7.38 | -3.724 | 0.001 | *** |
| STAI-state (/60) | 37.79 ± 10.411 | 27.79 ± 7.73 | 3.481 | 0.001 | *** |
| HAD-Depression (/21) | 5.63 ± 3.88 | 3.74 ± 2.84 | 2.825 | 0.005 | *** |
| HAD-Anxiety (/21) | 6.79 ± 3.75 | 5.32 ± 2.75 | 1.799 | 0.072 | ns |
| Starkstein apathy scale (/42) | 11.21 ± 4.2 | 8.67 ± 5.30 | 2.068 | 0.041 | ns |
| MDS-UPDRS III (/132) | 29.66 ± 13.80 | 11.19 ± 7.62 | 3.34 | 0.001 | *** |
| Dyskinesia scale (/28) | 0 ± 0 | 3.95 ± 3.47 | NC | NC | NC |

### 8. Correlation between motor and neuropsychiatric transition dynamics: Supplementary Table 3

Correlations between transition timepoints of neuropsychiatric (NFS-ON and NFS-OFF item sub-scores) and motor (akinesia, rigidity) measures during the OFF-to-ON levodopa transition.

Transition timepoints (t25, t50, t75, t100) correspond to the time required to reach 25%, 50%, 75%, and 100% of the maximal response observed during the session.

Values are reported as Pearson’s correlation coefficients (R), with corresponding Fisher’s Z-transformed values, frequentist p-values, Bayes factors (BF), and FDR-corrected p-values.

Bayes factors quantify evidence for the presence (H1) or absence (H0) of an association. BF > 1 supports low evidence for an association between the scores, BF > 3 indicates moderate evidence for an association. BF < 1 favors the absence of association between the scores, BF < 1/3 indicates moderate evidence for absence of association. Correlations with BF > 1 are shown in **bold**, those with BF < 1 are shown in italic, and those with BF < 0.33 are shown in bold and italic.

|  |  | n eligible pairs | Pearson's R  Or Spearman’s Rho [#] (Fisher's Z) | p-value (BF) | FDR-corrected p-value |
| --- | --- | --- | --- | --- | --- |
| NFS-ON vs  NFS-OFF | Onset (t=25% max) | 18 | 0.29 (0.30) | 0.23 *(0.56)* | 0.6 |
|  | T50 (t=50% max) | 18 | 0.28 (0.29) | 0.26 *(0.53)* | 0.6 |
|  | T75 (t=75% max) | 18 | 0.44 (0.48) | 0.065(**1.41**) | 0.6 |
|  | T100 Stabilization (t=100% max) | 18 | 0.63 (0.74) | 0.005 (**11.41**)** | 0.10 |
| NFS-ON vs Akinesia | Onset (t=25% max) | 18 | 0.14 (0.14) | 0.57 (*0.34*) | 0.79 |
|  | T50 (t=50% max) | 19 | 0.22 (0.22) | 0.37 (*0.41*) | 0.6 |
|  | T75 (t=75% max) | 19 | 0.25 (0.25) | 0.30 (*0.46*) | 0.6 |
|  | T100 Stabilization (t=100% max) | 19 | 0.13 (0.13) | 0.59 (***0.33***) | 0.79 |
| NFS-ON vs Rigidity | Onset (t=25% max) | 15 | 0.28 (0.29) | 0.31 (*0.51*) | 0.6 |
|  | T50 (t=50% max) | 15 | 0.28 (0.29) | 0.31 (*0.51*) | 0.6 |
|  | T75 (t=75% max) | 15 | 0.34 (0.35) | 0.22 (*0.63*) | 0.6 |
|  | T100 Stabilization (t=100% max) | 15 | 0.24 (0.24) | 0.39 (*0.45*) | 0.6 |
| NFS-OFF vs Akinesia | Onset (t=25% max) | 17 | 0.015 # (0.015) | 0.955 (***0.312***) | 0.99 |
|  | T50 (t=50% max) | 18 | -0.08 (-0.08) | 0.76 **(*0.30***) | 0.95 |
|  | T75 (t=75% max) | 18 | 0.01 (0.01) | 0.99 (***0.29***) | 0.99 |
|  | T100 Stabilization (t=100% max) | 18 | 0.28 (0.29) | 0.26 (*0.52*) | 0.6 |
| NFS-OFF vs Rigidity | Onset (t=25% max) | 14 | 0.05 (0.05) | 0.86 (***0.33***) | 0.96 |
|  | T50 (t=50% max) | 14 | 0.06 (0.06) | 0.83 (*0.34*) | 0.96 |
|  | T75 (t=75% max) | 14 | 0.28 (0.29) | 0.33 (*0.51*) | 0.6 |
|  | T100 Stabilization (t=100% max) | 14 | 0.25 (0.26) | 0.38 (*0.47*) | 0.6 |

### 9. Association between transition dynamics and clinical profiles

To investigate whether interindividual differences in transition dynamics were associated with clinical and neuropsychological profiles, we performed correlation analyses between clinical scores and temporal delays across domains.

For each patient, transition delays were defined as the differences between motor (akinesia) and neuropsychiatric (NFS-ON and NFS-OFF) transition timepoints (t25, t50, t75, t100). For example, the delay between NFS-ON and NFS-OFF onset was computed as: NFS-ON t25 − NFS-OFF t25.

Associations between these delays and clinical/neuropsychological scores were assessed using both frequentist and Bayesian correlation analyses (JASP software). Pearson correlations were applied when normality assumptions were met (Shapiro-Wilk test), and Spearman rank correlations otherwise (materialized by #).

No associations between clinical scores and transition delays remained significant after FDR correction in the frequentist analysis. However, exploratory Bayesian analyses suggested potential associations between specific behavioral dimensions and transition asynchrony (see Supplementary Table 4).

### 10. Mean transition timepoints across clusters: Supplementary Table 4

Mean ± standard deviation (minutes) of transition timepoints (t25, t50, t75, t100) for motor and neuropsychiatric measures across identified clusters.

NA : rigidity score computed for less than 2 patients

|  |  | NFS-ON | NFS-OFF | Akinesia | Rigidity |
| --- | --- | --- | --- | --- | --- |
| C0: Early neuropsychiatric ON Transition | t25 | 8±4 | 4±1 | 19±7 | 15±8 |
|  | t50 | 12±5 | 7±2 | 27±5 | 26±6 |
|  | t75 | 16±6 | 10±3 | 33±5 | 32±8 |
|  | t100 | 24±9 | 22±11 | 39±6 | 46±15 |
| C1: Early neuropsychiatric OFF Transition | t25 | 24±1 | 9±7 | 24±11 | 23±10 |
|  | t50 | 31±4 | 14±6 | 37±1 | 33±4 |
|  | t75 | 38±5 | 27±12 | 42±1 | 40±1 |
|  | t100 | 47±6 | 43±14 | 46±4 | 46±4 |
| C3: Early Motor Transition | t25 | 30±16 | 22±9 | 12±4 | NA |
|  | t50 | 46±9 | 27±8 | 18±4 | NA |
|  | t75 | 53±9 | 43±14 | 24±2 | NA |
|  | t100 | 64±3 | 63±3 | 40±7 | NA |
| C4: Late-Co-Transition | t25 | 32±5 | 30±9 | 27±4 | 23±11 |
|  | t50 | 38±5 | 39±5 | 36±3 | 37±5 |
|  | t75 | 45±4 | 48±5 | 42±5 | 46±4 |
|  | t100 | 57±7 | 56±5 | 56±10 | 57±5 |
| C2: Early Co-transition | t25 | 20±6 | 22±7 | 17±12 | NA |
|  | t50 | 24±4 | 24±7 | 26±5 | NA |
|  | t75 | 31±2 | 34±4 | 36±2 | NA |
|  | t100 | 38±9 | 58±4 | 58±3 | NA |

### 11. Individual transition profiles across clusters: ****Supplementary Figure 2****

These profiles illustrate the diversity of temporal dynamics across patients, including synchronous transitions, earlier neuropsychiatric changes, and earlier motor responses.

Individual motor and neuropsychiatric transition profiles during the OFF-to-ON levodopa states, grouped according to cluster membership. For each patient, time courses of normalized scores are displayed for: neuropsychiatric symptoms NFS-ON (red), neuropsychiatric symptoms NFS-OFF (blue), akinesia (black), and rigidity (gray, when available). Scores are expressed as relative changes from the maximal OFF-state value. Dots represent observed values at each timepoint, while solid lines correspond to spline-interpolated curves used to estimate continuous transition dynamics. Horizontal dashed lines indicate transition thresholds corresponding to 25%, 50%, and 100% of the maximal response (t25, t50, t100). The time at which each curve crosses these thresholds defines the corresponding transition timepoints.

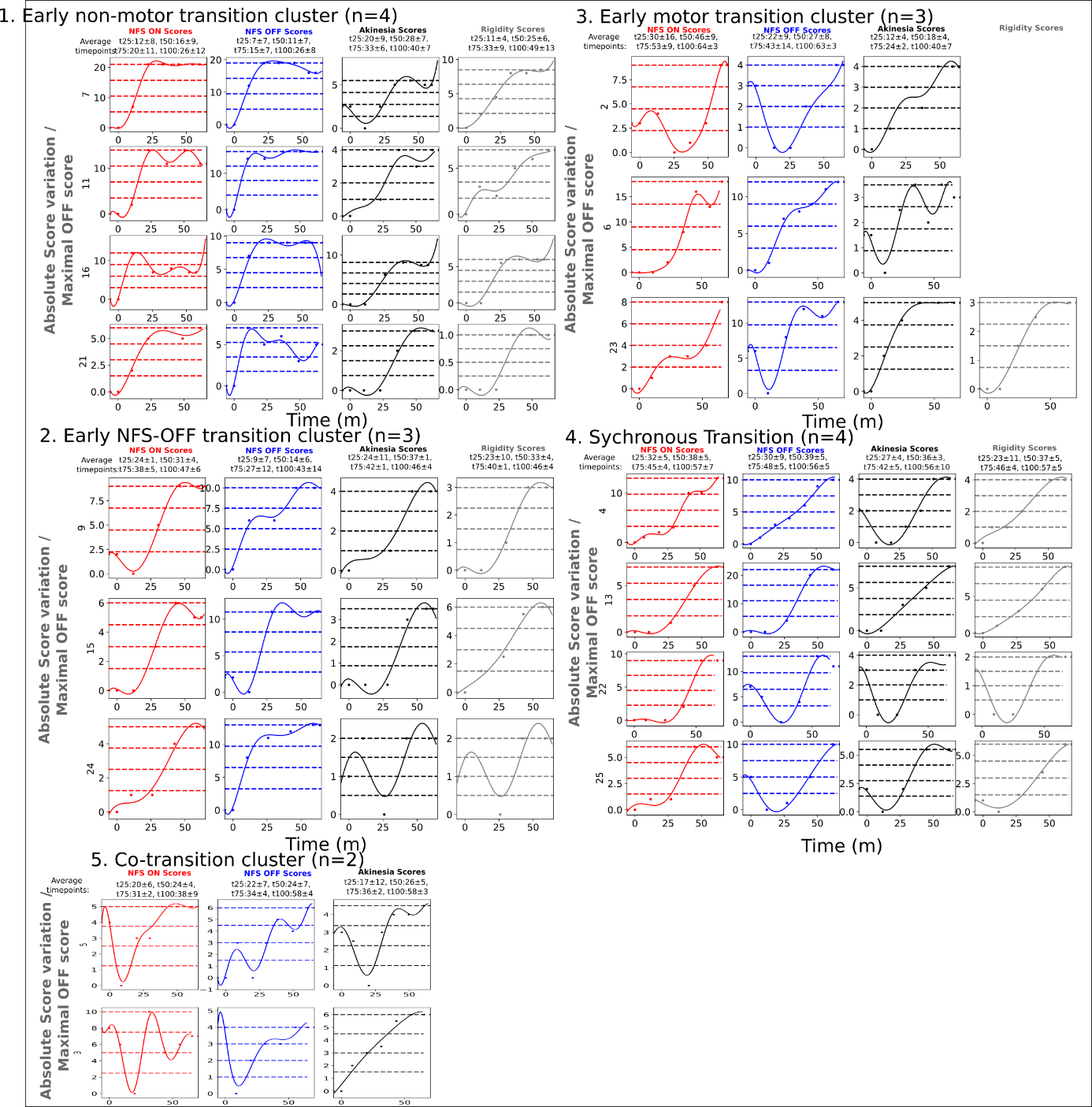

### 12. Individual transition profiles across clusters: ****Supplementary Figure 3****

Mean normalized trajectories of neuropsychiatric and motor scores during the 60 minutes following levodopa intake, grouped according to unsupervised clustering of transition times. Three main patterns were identified: synchronous transitions**,** earlier neuropsychiatric changes**,** and earlier motor responses. The most representative cluster for this pattern has been selected for the figure. Curves represent mean values, with shaded areas indicating standard deviations. OFF-related neuropsychiatric scores are inverted for visualization. Transition timepoints range (mean +-SE) are indicated by horizontal bars below each curve.

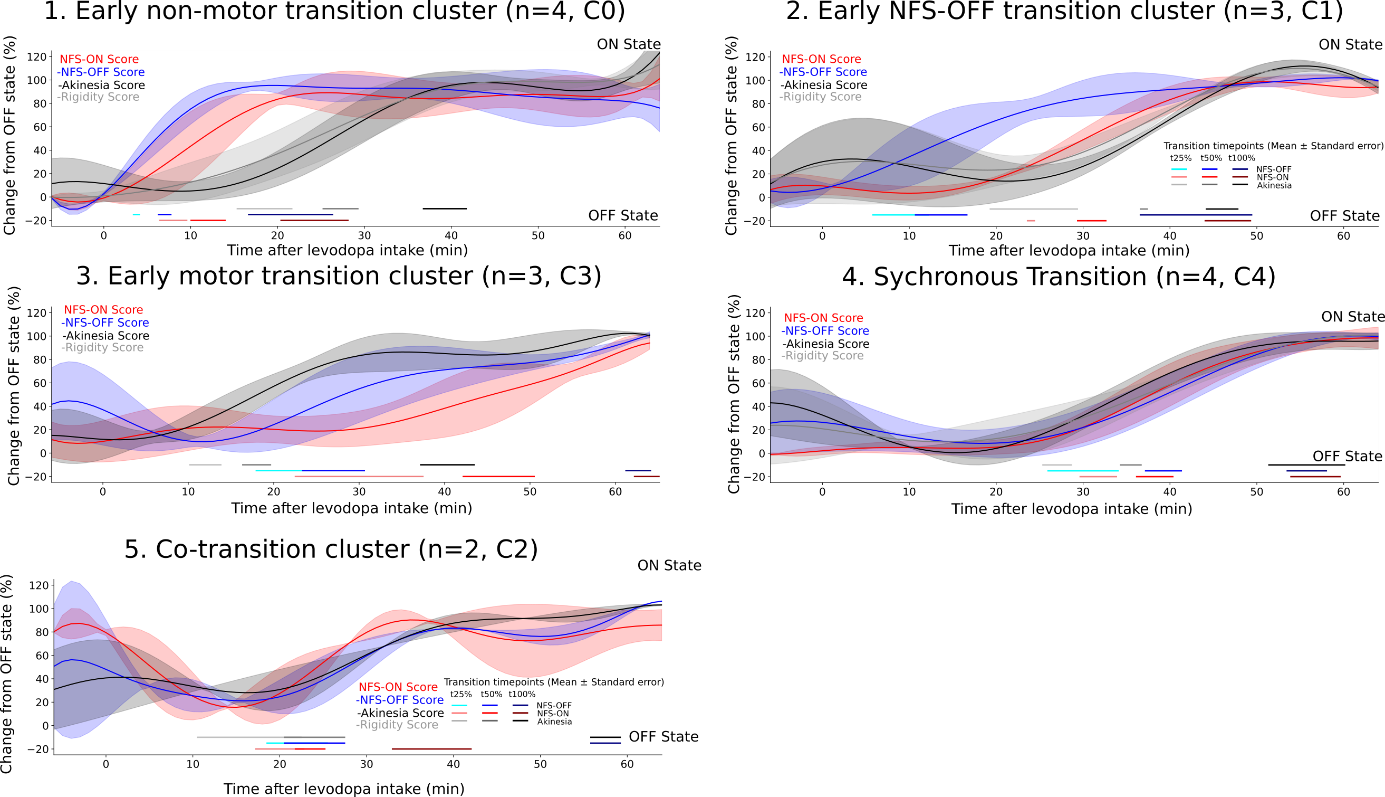
